# Quantifying Device Type and Handedness Biases in a Remote Parkinson’s Disease AI-Powered Assessment

**DOI:** 10.1101/2025.01.03.25319964

**Authors:** Zerin Nasrin Tumpa, Md Rahat Shahriar Zawad, Lydia Sollis, Shubham Parab, Irene Chen, Peter Washington

## Abstract

Early detection of Parkinson’s Disease (PD) can enable early access to care, improving patient outcomes. We investigate the use of machine learning to predict PD using data recorded from a web application measuring structured mouse and keypress data through tests assessing finger and hand movement patterns. We evaluate the impact of demographic bias and bias related to device type and handedness, which are particularly relevant to our application. We collected data from 251 participants (99 PD, 152 Non-PD). Using a random forest model, we observed an 84% F1 score, 86% sensitivity, and 92% specificity. When examining only F1-score differences across groups, no significant bias appears. However, conducting a more in-depth analysis using algorithmic fairness metrics uncovers bias regarding the positive prediction and error rates. In particular, we found that sex and ethnicity have no statistically significant impact on receiving a PD prediction. However, we observe biases regarding device type and dominant hand in terms of receiving a PD diagnosis, as evidenced by disparate impact and equalized odds fairness metrics. This work demonstrates that remote digital health diagnostics using consumer devices like desktops or laptops can exhibit nontraditional yet significant biases concerning understudied factors in algorithmic fairness such as device type and handedness.

## Introduction

PD is the fastest-growing neurological disorder in the world. It is projected that the number of individuals aged 50 years or older with PD in the USA will rise by 128 percent from about 340,000 in 2005 to around 610,000 by 2030^2^. Numerous studies have emphasized the possibility of diagnosing PD through motor patterns such as verbal fluency^3^, voice signals-voice disorders^4,5^, speech^6^, handwritten trace^7^, and gait patterns^8^. Finger tapping is one of the most reliable tests for motor performance ^9^. The necessity for lab equipment and specialist supervision has made the detection process more complex and less cost-effective^10^. We, therefore, aim to develop a flexible and cost-effective method for detecting PD using machine learning applied to mouse/trackpad and keyboard data. Our pilot study^11^ explored the feasibility of an earlier version of our web application, which demonstrated the potential for remote PD disease screening using digital motor assessments. Our current study builds upon these findings by refining the data collection framework, expanding participant recruitment from a small sample to over 250 participants, and systematically evaluating bias with respect to several factors.

We collected data from participants who engaged in structured interactive tasks on a remotely accessible web application that we created. Each assessment consisted of a series of mouse tracing and keyboard pressing tasks to measure motor movement. For the key press tasks, metrics such as response time, accuracy, and accidental key presses from unintended movements were recorded. For the mouse trace tasks, where participants were asked to trace straight and curved paths displayed on-screen using a trackpad or mouse, we measured the stability and precision of two-dimensional hand movements, providing insights into motor control and coordination of the lower arm.

While prior studies have explored remote digital health assessments for PD^12–17^, our study delves into issues of algorithmic fairness in remote PD assessments in particular and consumer digital health informatics more broadly by exploring the performance of machine learning models across diverse demographic groups in a web application for remote PD assessment. By examining factors such as sex, race, device type, and dominant hand using multiple fairness metrics, we aim to understand and highlight potential biases in the system’s predictions. We note in particular that exploring algorithmic fairness with respect to device type and dominant hand are understudied aspects of consumer digital health informatics.

## Results

### Novel Remotely Curated Dataset

We recruited 251 individuals, providing a comprehensive dataset to analyze the effectiveness of our remotely accessible web-based application for detecting PD. Of these participants, 99 were diagnosed with PD and 152 were Non-PD controls (Figure 1a).

**Figure 1:**
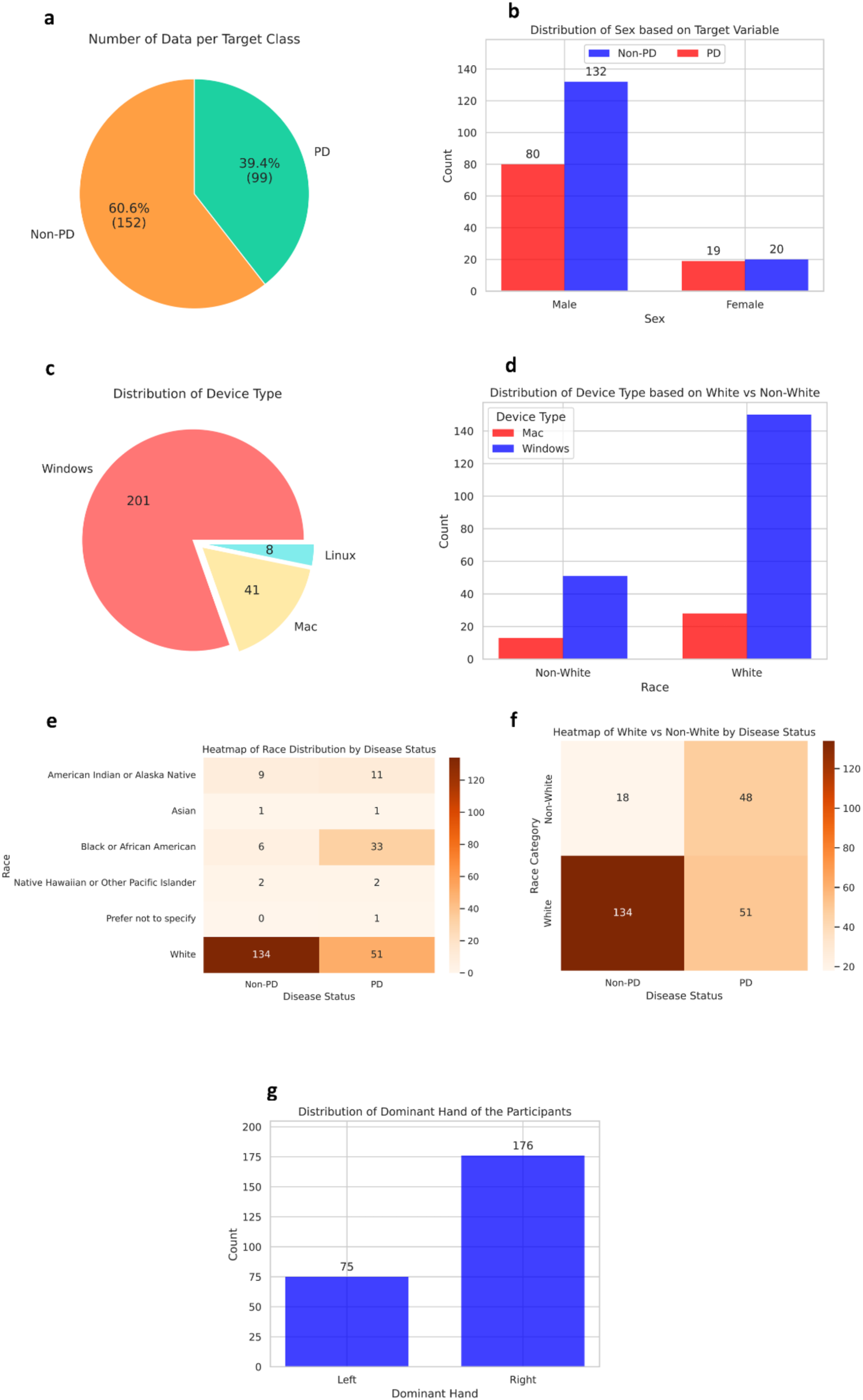
Data distributions before any upsampling. (a) The dataset is class imbalanced, with approximately 39.4% PD patients and 60.6% Non-PD participants. (b) The total number of male participants is much higher than that of female participants, with differing class balances across sex groups. (c) Most of the participants used a Windows device. Two hundred-one participants used Windows, whereas only 41 users used a Mac, and eight used Linux. (d) Within the White group, Windows is the predominant device type, with 150 users, compared to only 28 users on Mac. Similarly, in the Non-White group, Windows usage remains higher, with 45 users, while Mac usage is much lower, at 12 users. This pattern indicates a strong preference for Windows devices across racial groups, although the White group has a significantly larger sample size overall. (e) Of participants with PD, 51 are White, followed by Black or African American, with 33 responses. White participants consist of 134 responses in the Non-PD group, with only six responses from Black or African American participants. (f) Distribution of race by disease status, specifically comparing White and Non-White groups.

There was significant underrepresentation of many groups in our dataset, and we sought to study the effect of this imbalance on disparities in performance across groups. The sex distribution included 39 females and 212 males (Figure 1b). Participants utilized various devices to access the web-based application: 201 used Windows desktops, 41 used Mac desktops, and 8 used Linux desktops (Figure 1c). The device type distribution was roughly similar between White and Non-White groups (Figure 1d). The heatmap in Figure 1e shows how individuals are distributed across different racial categories based on their PD status. The majority of participants were White (185). There were 39 Black or African American participants. Smaller groups included 20 American Indian or Alaska Native individuals, 4 Native Hawaiian or other Pacific Islander individuals, and 2 Asian participants. While the distribution of White and Non-White groups was balanced within the PD group, the Non-PD group was heavily skewed towards White participants (Figure 1f). Most of the participants were right-handed, with 176 individuals identifying as right-handed and seventy-five as left-handed (Figure 1g).

The remotely accessible application included seven structured tasks: three involving mouse exercises, three involving keyboard interactions, and one testing working memory. Each task consisted of three levels of increasing difficulty. The mouse-based tasks involved tracing straight lines and spirals, while the keyboard-based tasks included single-letter presses, multiple-letter presses, and randomized sequences, forming a comprehensive set of activities designed to test different motor and cognitive skills. The working memory game assessed cognitive function. From these interactions, we extracted 80 features across five major categories.

### Model Selection

We first sought to select high-performing models for subsequent analyses. Using manually engineered features, we tested six classical machine-learning models for predicting PD. Each model was evaluated using 5-fold cross-validation on the training set, and the performance of the models was recorded using the mean F1 score across five folds (Figure 2). The dataset in each fold was split into 70% training and 30% testing data.

**Figure 2:**
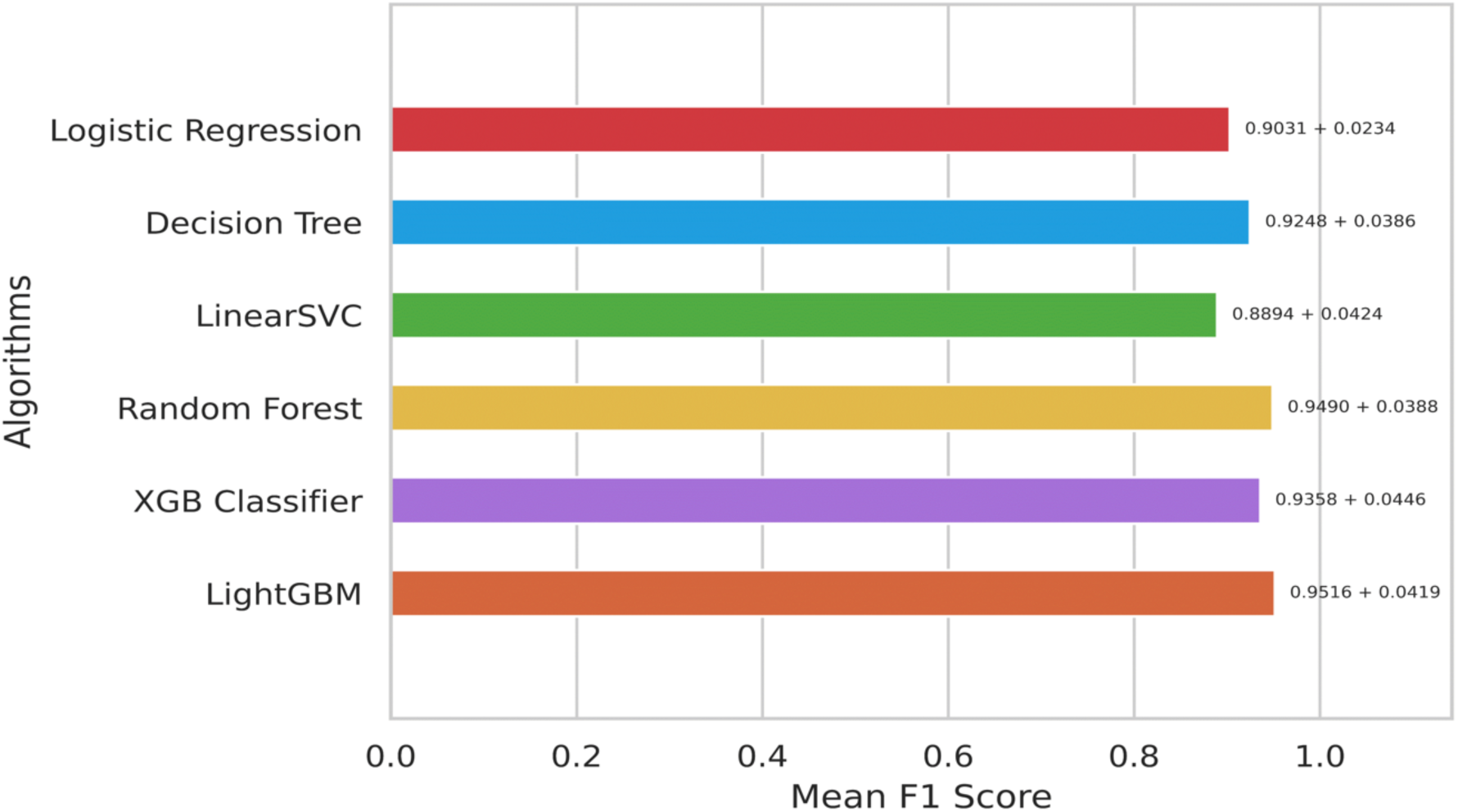
Comparison of classical machine learning models. The results are generated by performing cross-validation on our training data only. We report the mean with standard deviation as error bars across all the folds.

Although all the experimental models exhibited promising performance during this model section stage, the LightGBM classifier outperformed the others by a small margin, achieving a 95.2% F1 score with a 4.19% standard deviation.The Random Forest, XGBoost, and LightGBM classifiers were selected to test their performance further on held-out test data. The 30% test data were used to make predictions using the three trained models (Table 1, Figure 3).

**Table 1:** Performance of the top-performing ML models on held-out test data.

|  | F1 Score | Sensitivity | Specificity |
| --- | --- | --- | --- |
| Random Forest | 84% | 86% | 92% |
| LightGBM | 78% | 78% | 90% |
| XGBoost | 77% | 81% | 88% |

**Figure 3:**
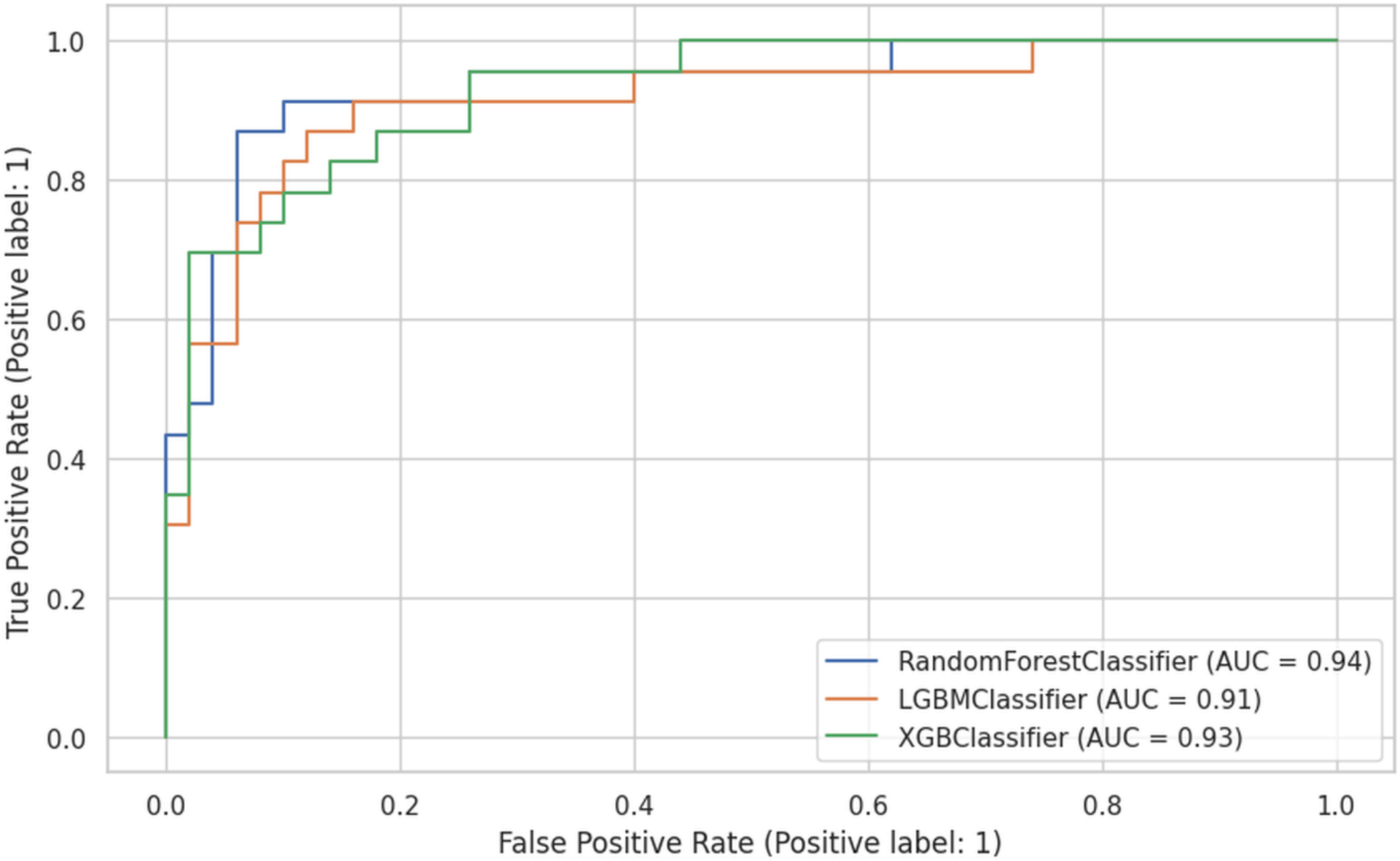
ROC curves and corresponding AUC scores for the random forest, LightGBM, and XGBoost classifiers. During the training phase, the three top-performing models were further tested on the held-out data to report their ROC curve and AUROC scores. The performance differences are minimal when comparing the ROC curves and the area under the curve (AUC). The Random Forest model achieved the highest AUC score of 94%, followed by XGBoost at 93% and LightGBM at 91%.

### Model Bias and Fairness

To evaluate bias, we used traditional accuracy metrics like F1-score, sensitivity, specificity, and AUC as well as fairness-specific metrics such as Disparate Impact, Equal Opportunity, and Equalized Odds. Because we measured these metrics as a ratio across groups, a value of 1 for these fairness metrics indicates perfect fairness, with equal treatment of the privileged and unprivileged groups. Values less than 1 signify biases against the unprivileged group, while values greater than 1 indicate potential biases favoring the unprivileged group.

We selected male, White, Mac, and right-handed individuals as the privileged groups in the study (Table 2). While the selection of unprivileged groups corresponded to underrepresentation in our dataset in most cases, we chose to set Mac users as the privileged group despite underrepresentation due to the homogeneity of Mac devices – a contrast to Windows devices created by a wide range of manufacturers.

**Table 2:** Our selection of “Privileged” and “Unprivileged” groups in terms of the calculations for common algorithmic fairness metrics.

| Category | Unprivileged | Privileged |
| --- | --- | --- |
| Sex | Female | Male |
| Race | Non-White (Black, Asian, Native Hawaiian etc.) | White |
| Device | Windows | Mac |
| Dominant Hand | Left | Right |

Tables 3.1 and 3.2 display the F1-score, sensitivity, specificity, specificity, and AUROC differences between groups for four attributes: sex, race, device type, and dominant hand. Table 3.1 shows these metrics following upsampling by race, while Table 3.2 provides the values before any resampling was applied.

**Table 3.1:**
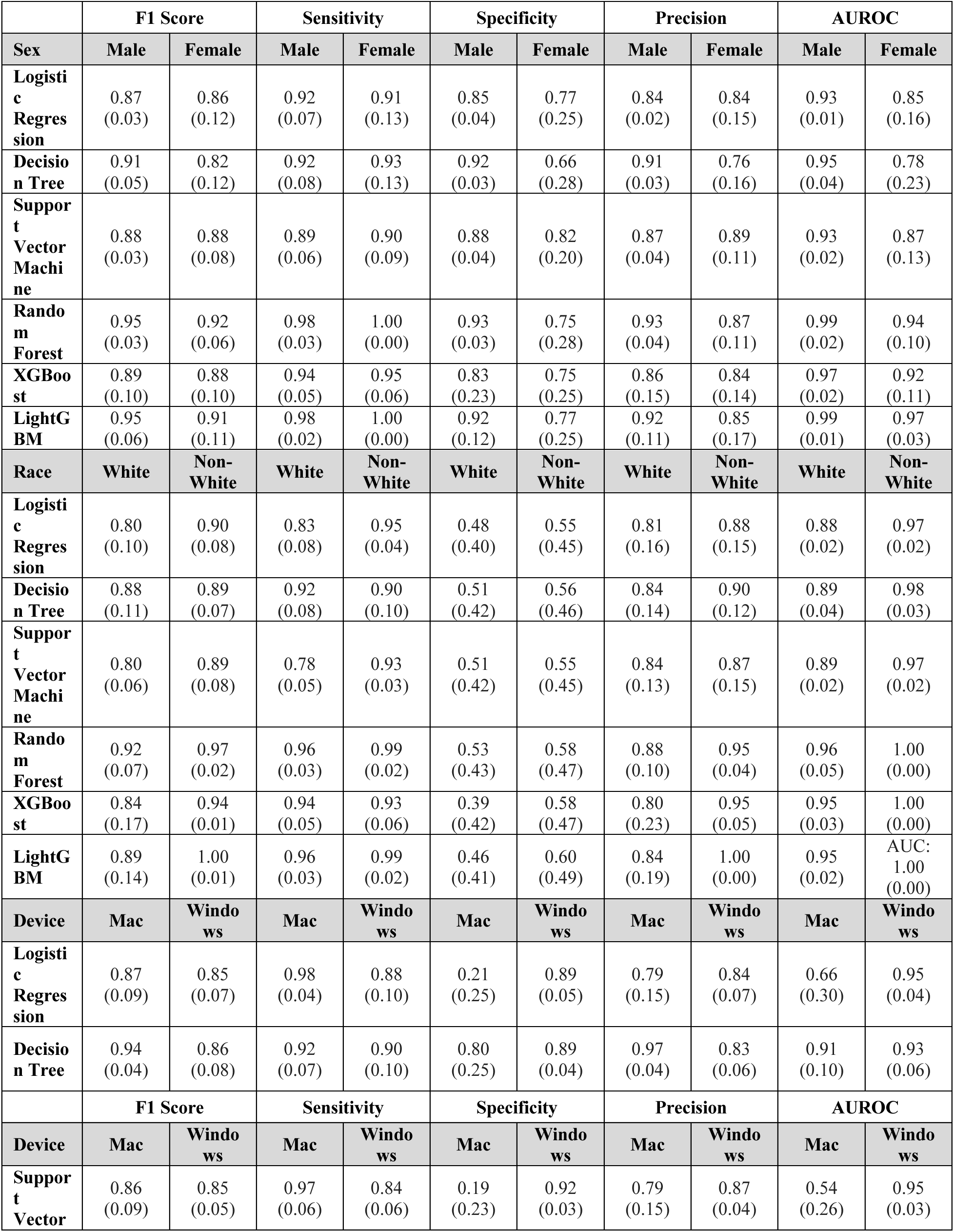
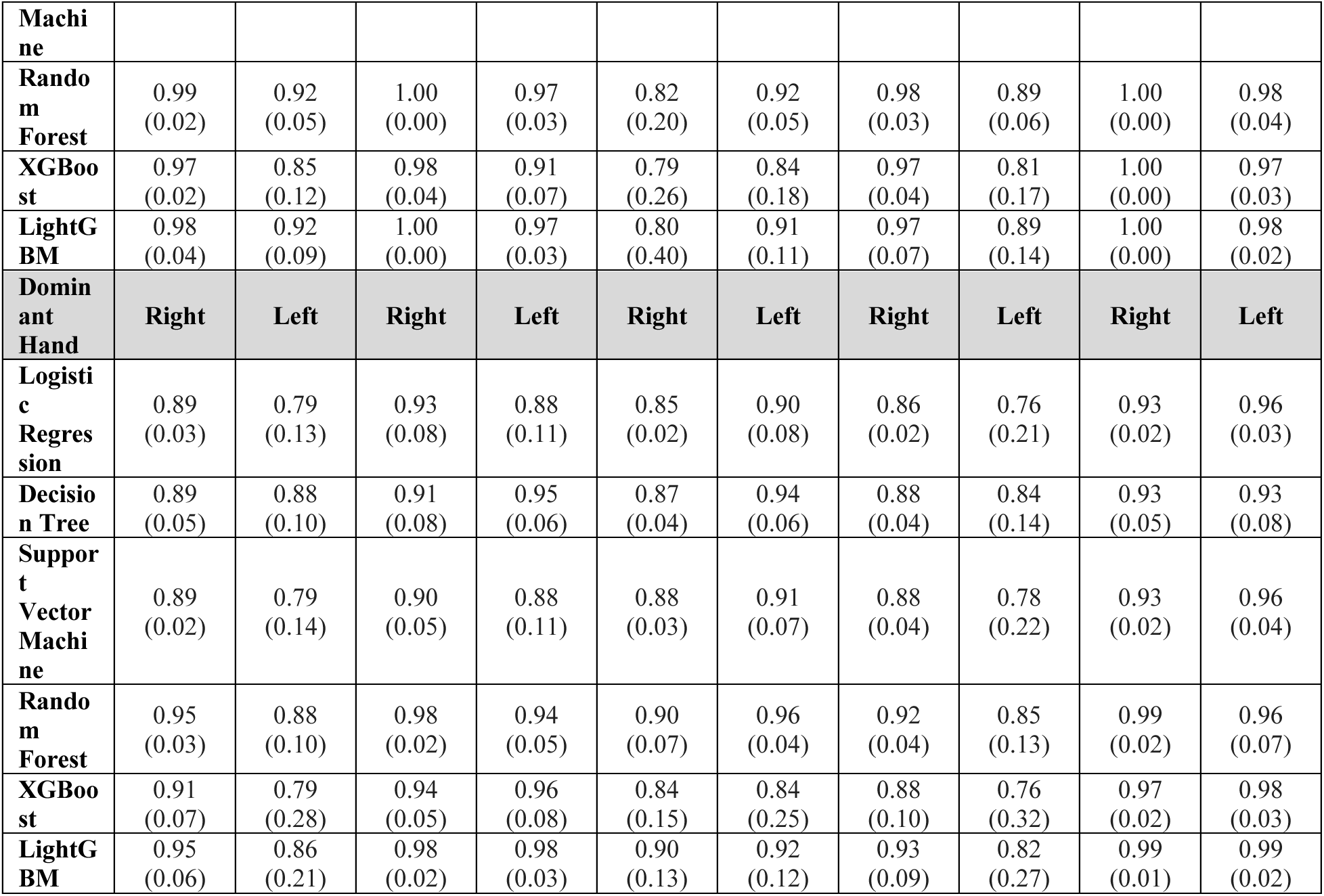
Model performance *after* race upsampling on each group based on four protected attributes: sex, race, device, and dominant hand.

**Table 3.2:** Model performance *before* race upsampling on each group based on four protected attributes: Sex, race, device, and dominant hand.

|  | **F1 Score** |  | **Sensitivity** |  | **Specificity** |  | **Precision** |  | **AUROC** |  |
| --- | --- | --- | --- | --- | --- | --- | --- | --- | --- | --- |
| **Sex** | **Male** | **Female** | **Male** | **Female** | **Male** | **Female** | **Male** | **Female** | **Male** | **Female** |
| **Logistic Regression** | 0.87<br>(0.03) | 0.80<br>(0.18) | 0.88<br>(0.10) | 0.90<br>(0.13) | 0.89<br>(0.06) | 0.67<br>(0.31) | 0.87<br>(0.07) | 0.80<br>(0.25) | 0.93<br>(0.04) | 0.76<br>(0.19) |
| **Decision Tree** | 0.82<br>(0.07) | 0.67<br>(0.22) | 0.84<br>(0.12) | 0.76<br>(0.27) | 0.84<br>(0.06) | 0.55<br>(0.28) | 0.82<br>(0.05) | 0.69<br>(0.24) | 0.85<br>(0.07) | 0.74<br>(0.12) |
| **Support Vector Machine** | 0.87<br>(0.03) | 0.80<br>(0.18) | 0.89<br>(0.09) | 0.85<br>(0.20) | 0.89<br>(0.05) | 0.81<br>(0.25) | 0.87<br>(0.05) | 0.84<br>(0.24) | 0.93<br>(0.04) | 0.78<br>(0.24) |
| **Random Forest** | 0.93<br>(0.03) | 0.83<br>(0.15) | 0.95<br>(0.05) | 0.97<br>(0.06) | 0.92<br>(0.05) | 0.62<br>(0.21) | 0.91<br>(0.06) | 0.76<br>(0.21) | 0.97<br>(0.03) | 0.94<br>(0.08) |
| **XGBoost** | 0.82<br>(0.12) | 0.79<br>(0.13) | 0.93<br>(0.07) | 0.97<br>(0.06) | 0.69<br>(0.35) | 0.58<br>(0.13) | 0.77<br>(0.20) | 0.69<br>(0.18) | 0.96<br>(0.03) | 0.90<br>(0.09) |
| **LightGBM** | 0.88<br>(0.04) | 0.85<br>(0.11) | 0.91<br>(0.08) | 0.93<br>(0.08) | 0.88<br>(0.08) | 0.71<br>(0.17) | 0.87<br>(0.08) | 0.80<br>(0.16) | 0.96<br>(0.05) | 0.92<br>(0.08) |
| **Race** | **White** | **Non-White** | **White** | **Non-White** | **White** | **Non-White** | **White** | **Non-White** | **White** | **Non-White** |
| **Logistic** | 0.71<br>(0.14) | 0.92<br>(0.06) | 0.74<br>(0.12) | 0.97<br>(0.03) | 0.92<br>(0.07) | 0.27<br>(0.39) | 0.77<br>(0.23) | 0.89<br>(0.11) | 0.92<br>(0.02) | 0.88<br>(0.09) |
| **Regression** |  |  |  |  |  |  |  |  |  |  |
| **Decision Tree** | 0.68<br>(0.10) | 0.86<br>(0.06) | 0.80<br>(0.15) | 0.82<br>(0.07) | 0.82<br>(0.05) | 0.56<br>(0.33) | 0.64<br>(0.14) | 0.91<br>(0.09) | 0.83<br>(0.08) | 0.70<br>(0.06) |
| **Support Vector Machine** | 0.73<br>(0.09) | 0.93<br>(0.05) | 0.75<br>(0.12) | 0.97<br>(0.03) | 0.92<br>(0.04) | 0.29<br>(0.40) | 0.77<br>(0.14) | 0.90<br>(0.09) | 0.91<br>(0.01) | 0.87<br>(0.12) |
| **Random Forest** | 0.84<br>(0.11) | 0.94<br>(0.04) | 0.93<br>(0.04) | 0.98<br>(0.03) | 0.92<br>(0.06) | 0.44<br>(0.39) | 0.80<br>(0.18) | 0.91<br>(0.09) | 0.97<br>(0.02) | 0.86<br>(0.11) |
| **XGBoost** | 0.71<br>(0.23) | 0.89<br>(0.09) | 0.94<br>(0.07) | 0.90<br>(0.06) | 0.71<br>(0.31) | 0.46<br>(0.44) | 0.65<br>(0.30) | 0.90<br>(0.13) | 0.95<br>(0.02) | 0.91<br>(0.05) |
| **LightGBM** | 0.79<br>(0.11) | 0.91<br>(0.06) | 0.88<br>(0.07) | 0.95<br>(0.05) | 0.91<br>(0.07) | 0.41<br>(0.40) | 0.78<br>(0.20) | 0.90<br>(0.11) | 0.94<br>(0.04) | 0.94<br>(0.04) |
| **Device** | **Mac** | **Windows** | **Mac** | **Windows** | **Mac** | **Windows** | **Mac** | **Windows** | **Mac** | **Windows** |
| **Logistic Regression** | 0.95<br>(0.05) | 0.80<br>(0.05) | 1.00<br>(0.00) | 0.81<br>(0.13) | 0.16<br>(0.20) | 0.89<br>(0.08) | 0.91<br>(0.09) | 0.84<br>(0.10) | 0.72<br>(0.20) | 0.90<br>(0.03) |
| **Decision Tree** | 0.83<br>(0.19) | 0.75<br>(0.04) | 0.79<br>(0.26) | 0.81<br>(0.12) | 0.43<br>(0.39) | 0.80<br>(0.07) | 0.96<br>(0.05) | 0.72<br>(0.05) | 0.78<br>(0.05) | 0.85<br>(0.03) |
|  | **F1 Score** |  | **Sensitivity** |  | **Specificity** |  | **Precision** |  | **AUROC** |  |
| **Device** | **Mac** | **Windows** | **Mac** | **Windows** | **Mac** | **Windows** | **Mac** | **Windows** | **Mac** | **Windows** |
| **Support Vector Machine** | 0.94<br>(0.04) | 0.81<br>(0.06) | 0.98<br>(0.04) | 0.80<br>(0.13) | 0.15<br>(0.20) | 0.91<br>(0.04) | 0.91<br>(0.08) | 0.85<br>(0.05) | 0.51<br>(0.00) | 0.90<br>(0.04) |
| **Random Forest** | 0.98<br>(0.03) | 0.87<br>(0.04) | 1.00<br>(0.00) | 0.91<br>(0.06) | 0.43<br>(0.39) | 0.89<br>(0.08) | 0.96<br>(0.05) | 0.86<br>(0.08) | 0.83<br>(0.09) | 0.97<br>(0.04) |
| **XGBoost** | 0.96<br>(0.05) | 0.77<br>(0.12) | 0.99<br>(0.03) | 0.92<br>(0.08) | 0.33<br>(0.42) | 0.69<br>(0.30) | 0.94<br>(0.08) | 0.71<br>(0.20) | 0.97<br>(0.03) | 0.94<br>(0.02) |
| **LightGBM** | 0.96<br>(0.03) | 0.83<br>(0.03) | 0.96<br>(0.05) | 0.87<br>(0.08) | 0.44<br>(0.39) | 0.87<br>(0.07) | 0.96<br>(0.05) | 0.82<br>(0.08) | 0.92<br>(0.11) | 0.94<br>(0.04) |
| **Dominant Hand** | **Right** | **Left** | **Right** | **Left** | **Right** | **Left** | **Right** | **Left** | **Right** | **Left** |
| **Logistic Regression** | 0.85<br>(0.02) | 0.88<br>(0.06) | 0.88<br>(0.10) | 0.88<br>(0.09) | 0.80<br>(0.12) | 0.98<br>(0.03) | 0.85<br>(0.08) | 0.92<br>(0.13) | 0.90<br>(0.03) | 0.92<br>(0.04) |
| **Decision Tree** | 0.80<br>(0.05) | 0.64<br>(0.18) | 0.81<br>(0.10) | 0.73<br>(0.20) | 0.78<br>(0.08) | 0.85<br>(0.12) | 0.82<br>(0.05) | 0.66<br>(0.24) | 0.82<br>(0.03) | 0.77<br>(0.09) |
| **Support Vector Machine** | 0.86<br>(0.03) | 0.86<br>(0.07) | 0.88<br>(0.10) | 0.86<br>(0.10) | 0.84<br>(0.08) | 0.97<br>(0.04) | 0.87<br>(0.07) | 0.88<br>(0.12) | 0.90<br>(0.03) | 0.93<br>(0.03) |
| **Random Forest** | 0.91<br>(0.02) | 0.86<br>(0.14) | 0.94<br>(0.03) | 0.92<br>(0.08) | 0.85<br>(0.09) | 0.95<br>(0.07) | 0.89<br>(0.05) | 0.84<br>(0.20) | 0.97<br>(0.03) | 0.97<br>(0.03) |
| **XGBoost** | 0.84<br>(0.07) | 0.67<br>(0.32) | 0.92<br>(0.06) | 0.92<br>(0.10) | 0.67<br>(0.27) | 0.72<br>(0.35) | 0.80<br>(0.13) | 0.64<br>(0.38) | 0.93<br>(0.04) | 0.96<br>(0.03) |
| **LightGBM** | 0.88<br>(0.03) | 0.74<br>(0.17) | 0.90<br>(0.03) | 0.88<br>(0.10) | 0.84<br>(0.09) | 0.90<br>(0.10) | 0.88<br>(0.06) | 0.70<br>(0.25) | 0.93<br>(0.04) | 0.95<br>(0.04) |

Notably, an unusual result highlights how underrepresentation can mask underlying biases in machine learning. Before upsampling, the Non-White group exhibited higher F1 scores than the White group, which was counter-intuitive given the underrepresentation of Non-White data. Closer examination revealed that the “Non-White Non-PD” group had substantially less data than the “White Non-PD” group, likely resulting in an inflated F1 score for the Non-White group overall. Figure 1f illustrates the distribution of participants by race and disease status, clearly highlighting the underrepresentation of the "Non-White Non-PD" group. This is a salient example of how the underrepresentation of certain groups in machine-learning datasets can mask underlying biases.

To address this imbalance, we increased the data in the “Non-White Non-PD” group to bring it closer in size to the White group using resampling with replacement. After race resampling, we observed that the F1 score gap between White and Non-White groups significantly narrowed, with no significant differences between the groups after upsampling (Table 5.1). All sensitive categories no longer yielded statistically significant differences in F1 scores (Figure 4).

**Table 4.1:** Bias metrics of the models based on the demographic groups from the cross-validation with bootstrap sampling *after* race upsampling. All metrics are reported as ratios between the privileged and underprivileged groups.

| Sex |  |  |  |
| --- | --- | --- | --- |
|  | Disparate Impact | Equal Opportunity | Equalized Odds |
| Logistic Regression | 0.99 (0.55) | 0.99 (0.11) | 0.25 (0.17) |
| Decision Tree | 1.18 (0.59) | 1.02 (0.16) | 0.30 (0.23) |
| Support Vector Machine | 0.99 (0.57) | 1.01 (0.04) | 0.22 (0.12) |
| Random Forest | 1.07 (0.59) | 1.02 (0.03) | 0.23 (0.22) |
| XGBoost | 1.03 (0.59) | 1.02 (0.03) | 0.28 (0.21) |
| LightGBM | 1.09 (0.55) | 1.02 (0.03) | 0.17 (0.21) |
| Race |  |  |  |
|  | Disparate Impact | Equal Opportunity | Equalized Odds |
| Logistic Regression | 1.44 (1.13) | 1.17 (0.08) | 0.20 (0.05) |
| Decision Tree | 1.22 (0.87) | 0.99 (0.07) | 0.13 (0.05) |
| Support Vector Machine | 1.59 (1.28) | 1.22 (0.10) | 0.21 (0.03) |
| Random Forest | 1.41 (1.14) | 1.03 (0.03) | 0.09 (0.05) |
| XGBoost | 1.24 (1.14) | 0.99 (0.01) | 0.22 (0.29) |
| LightGBM | 1.44 (1.43) | 1.03 (0.03) | 0.15 (0.20) |
| Device |  |  |  |
|  | Disparate Impact | Equal Opportunity | Equalized Odds |
| Logistic Regression | 0.42 (0.03) | 0.90 (0.07) | 0.71 (0.27) |
| Decision Tree | 0.60 (0.15) | 0.99 (0.13) | 0.24 (0.15) |
| Support Vector Machine | 0.38 (0.04) | 0.87 (0.04) | 0.74 (0.24) |
| Random Forest | 0.48 (0.05) | 0.96 (0.03) | 0.15 (0.14) |
| XGBoost | 0.58 (0.14) | 0.94 (0.08) | 0.17 (0.15) |
| LightGBM | 0.54 (0.10) | 0.97 (0.03) | 0.19 (0.27) |
| Dominant Hand |  |  |  |
|  | Disparate Impact | Equal Opportunity | Equalized Odds |
| Logistic Regression | 0.55 (0.18) | 0.95 (0.10) | 0.15 (0.05) |
| Decision Tree | 0.57 (0.22) | 1.06 (0.04) | 0.11 (0.04) |
| Support Vector Machine | 0.57 (0.14) | 0.98 (0.12) | 0.16 (0.05) |
| Random Forest | 0.54 (0.22) | 0.96 (0.04) | 0.12 (0.06) |
| XGBoost | 0.70 (0.24) | 1.03 (0.10) | 0.12 (0.06) |
| LightGBM | 0.60 (0.18) | 1.01 (0.04) | 0.06 (0.05) |

**Table 4.2:** Bias metrics of the models based on the demographic groups from the cross-validation with bootstrap sampling *before* race upsampling. All metrics are reported as ratios between the privileged and underprivileged groups.

| Sex |  |  |  |
| --- | --- | --- | --- |
|  | Disparate Impact | Equal Opportunity | Equalized Odds |
| Logistic Regression | 1.42 (0.48) | 1.03 (0.14) | 0.34 (0.19) |
| Decision Tree | 1.39 (0.68) | 0.91 (0.32) | 0.46 (0.13) |
| Support Vector Machine | 1.21 (0.45) | 0.96 (0.20) | 0.29 (0.12) |
| Random Forest | 1.51 (0.39) | 1.03 (0.08) | 0.34 (0.14) |
| XGBoost | 1.37 (0.48) | 1.06 (0.09) | 0.34 (0.13) |
| LightGBM | 1.35 (0.42) | 1.04 (0.09) | 0.27 (0.08) |
| Race |  |  |  |
|  | Disparate Impact | Equal Opportunity | Equalized Odds |
| Logistic Regression | 3.55 (0.76) | 1.41 (0.25) | 0.57 (0.28) |
| Decision Tree | 2.29 (0.51) | 1.09 (0.19) | 0.24 (0.06) |
| Support Vector Machine | 3.83 (1.45) | 1.46 (0.33) | 0.57 (0.29) |
| Random Forest | 2.98 (0.87) | 1.06 (0.07) | 0.34 (0.29) |
| XGBoost | 1.93 (0.68) | 0.95 (0.09) | 0.47 (0.29) |
| LightGBM | 2.80 (0.48) | 1.10 (0.14) | 0.40 (0.29) |
| Device |  |  |  |
|  | Disparate Impact | Equal Opportunity | Equalized Odds |
| Logistic Regression | 0.39 (0.09) | 0.81 (0.13) | 0.52 (0.22) |
| Decision Tree | 0.73 (0.39) | 1.31 (0.83) | 0.33 (0.13) |
| Support Vector Machine | 0.38 (0.06) | 0.81 (0.11) | 0.49 (0.26) |
| Random Forest | 0.45 (0.05) | 0.91 (0.06) | 0.26 (0.12) |
| XGBoost | 0.58 (0.18) | 0.93 (0.07) | 0.23 (0.11) |
| LightGBM | 0.47 (0.03) | 0.91 (0.07) | 0.24 (0.08) |
| Dominant Hand |  |  |  |
|  | Disparate Impact | Equal Opportunity | Equalized Odds |
| Logistic Regression | 0.42 (0.18) | 1.01 (0.05) | 0.24 (0.03) |
| Decision Tree | 0.54 (0.20) | 0.92 (0.22) | 0.27 (0.07) |
| Support Vector Machine | 0.48 (0.21) | 1.00 (0.05) | 0.21 (0.04) |
| Random Forest | 0.49 (0.22) | 0.98 (0.09) | 0.17 (0.04) |
| **XGBoost** | 0.73 (0.18) | 1.02 (0.15) | 0.19 (0.06) |
| **LightGBM** | 0.55 (0.16) | 0.98 (0.10) | 0.19 (0.05) |

**Table 5.1:**
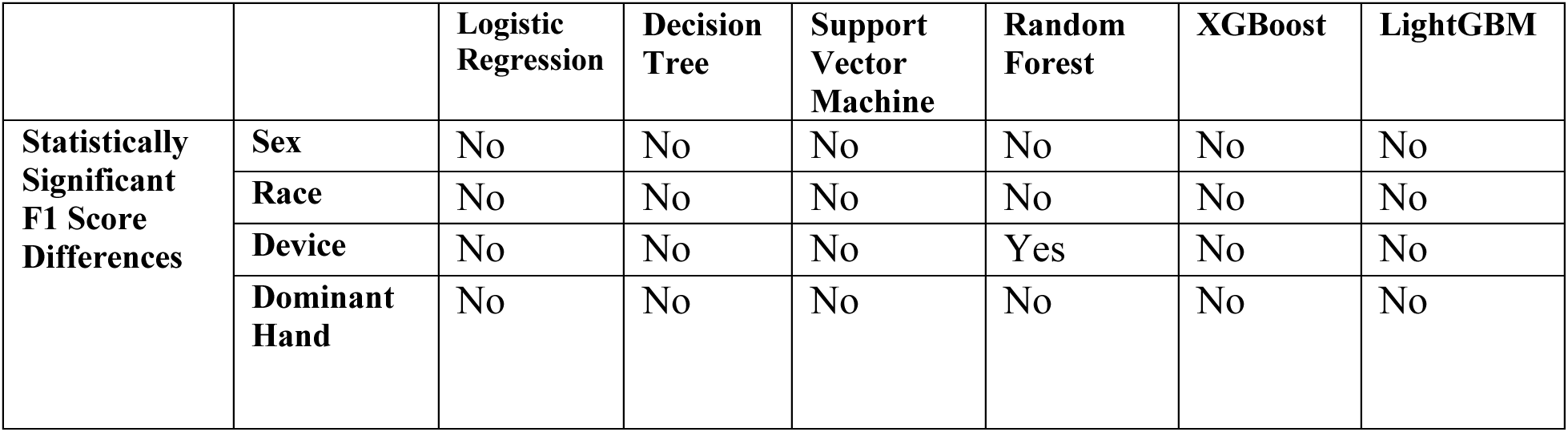
Binary indicator of whether a statistically significant difference in F1-score is observed using a t-test between groups for each sensitive attribute (*after* race upsampling).

|  |  | **Logistic Regression** | **Decision Tree** | **Support Vector Machine** | **Random Forest** | **XGBoost** | **LightGBM** |
| --- | --- | --- | --- | --- | --- | --- | --- |
| **Statistically Significant F1 Score Differences** | **Sex** | No | No | No | No | No | No |
|  | **Race** | No | No | No | No | No | No |
|  | **Device** | No | No | No | Yes | No | No |
|  | **Dominant Hand** | No | No | No | No | No | No |

**Table 5.2:**
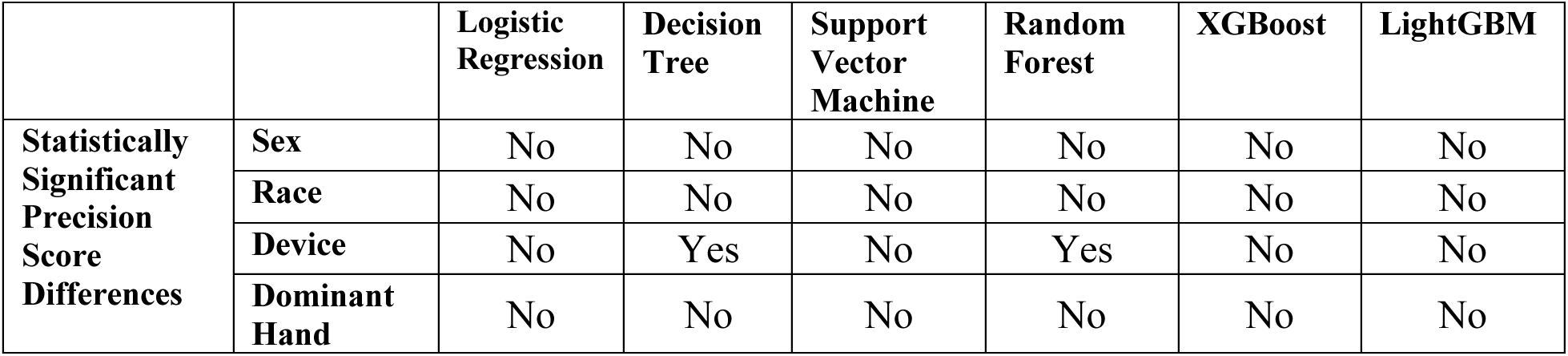
Binary indicator of whether a statistically significance difference in precision is observed using a t-test between groups for each sensitive attribute (*after* race upsampling).

|  |  | **Logistic Regression** | **Decision Tree** | **Support Vector Machine** | **Random Forest** | **XGBoost** | **LightGBM** |
| --- | --- | --- | --- | --- | --- | --- | --- |
| **Statistically Significant Precision Score Differences** | **Sex** | No | No | No | No | No | No |
|  | **Race** | No | No | No | No | No | No |
|  | **Device** | No | Yes | No | Yes | No | No |
|  | **Dominant Hand** | No | No | No | No | No | No |

**Table 5.3:**
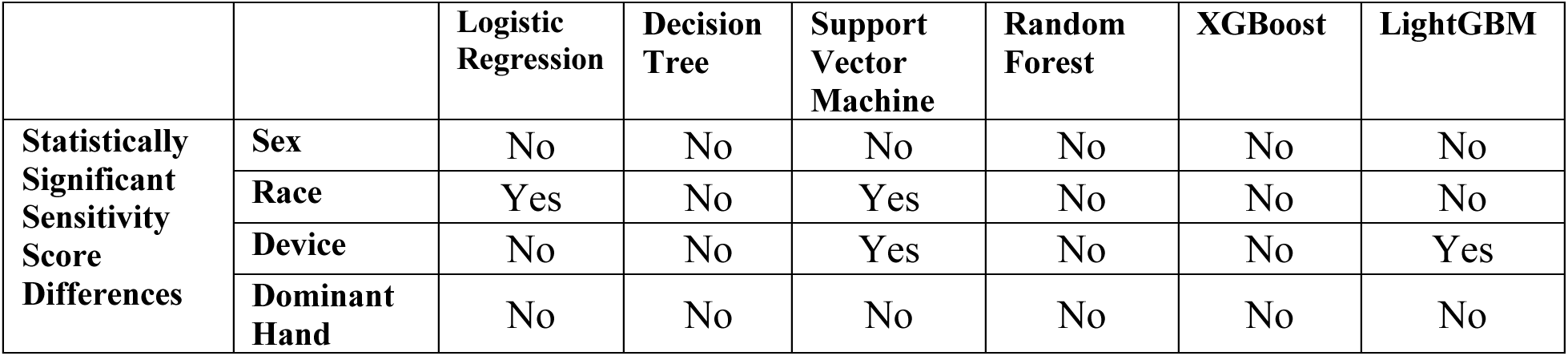
Binary indicator of whether a statistically significance difference in sensitivity is observed using a t-test between groups for each sensitive attribute (*after* race upsampling).

**Table 5.4:** Binary indicator of whether a statistically significance difference in specificity is observed using a t-test between groups for each sensitive attribute (*after* race upsampling).

|  |  | **Logistic Regression** | **Decision Tree** | **Support Vector Machine** | **Random Forest** | **XGBoost** | **LightGBM** |
| --- | --- | --- | --- | --- | --- | --- | --- |
| **Statistically Significant** | **Sex** | No | No | No | No | No | No |
|  | **Race** | Yes | No | Yes | No | No | No |
| **Specificity Score Differences** | **Device** | No | No | Yes | No | No | Yes |
|  | **Dominant Hand** | No | No | No | No | No | No |

**Table 5.5:** Binary indicator of whether a statistical difference in F1-score is observed using a t-test between groups for each sensitive attribute (*before* race upsampling).

|  |  | **Logistic Regression** | **Decision Tree** | **Support Vector Machine** | **Random Forest** | **XGBoost** | **LightGBM** |
| --- | --- | --- | --- | --- | --- | --- | --- |
| **Statistically Significant F1 Score Differences** | **Sex** | No | No | No | No | No | No |
|  | **Race** | Yes | Yes | Yes | No | No | No |
|  | **Device** | Yes | No | Yes | Yes | Yes | Yes |
|  | **Dominant Hand** | No | No | No | No | No | No |

**Table 5.6:** Binary indicator of whether a statistically significance difference in precision is observed using a t-test between groups for each sensitive attribute (*before* race upsampling).

|  |  | **Logistic Regression** | **Decision Tree** | **Support Vector Machine** | **Random Forest** | **XGBoost** | **LightGBM** |
| --- | --- | --- | --- | --- | --- | --- | --- |
| **Statistically Significant Precision Score Differences** | **Sex** | No | No | No | No | No | No |
|  | **Race** | No | No | No | No | No | No |
|  | **Device** | No | Yes | No | No | Yes | No |
|  | **Dominant Hand** | No | No | No | No | No | No |

**Table 5.7:** Binary indicator of whether a statistically significance difference in sensitivity is observed using a t-test between groups for each sensitive attribute (*before* race upsampling).

|  |  | **Logistic Regression** | **Decision Tree** | **Support Vector Machine** | **Random Forest** | **XGBoost** | **LightGBM** |
| --- | --- | --- | --- | --- | --- | --- | --- |
| **Statistically Significant Sensitivity Score Differences** | **Sex** | No | No | No | No | No | No |
|  | **Race** | Yes | No | Yes | No | No | No |
|  | **Device** | Yes | No | Yes | Yes | No | No |
|  | **Dominant Hand** | No | No | No | No | No | No |

**Table 5.8:**
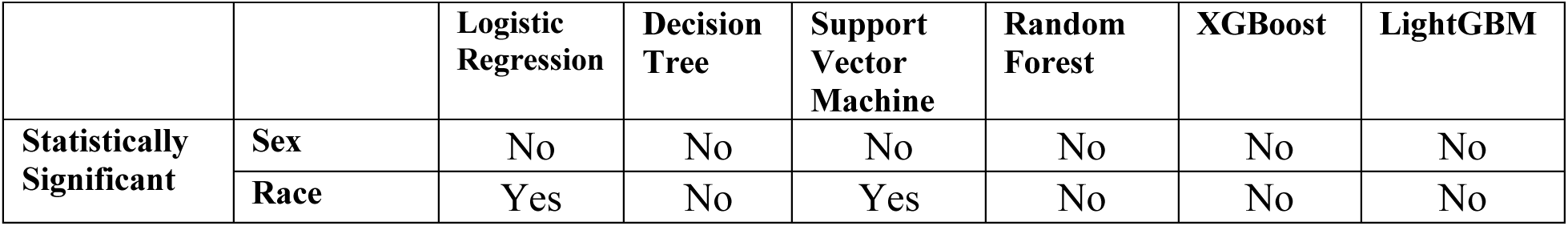
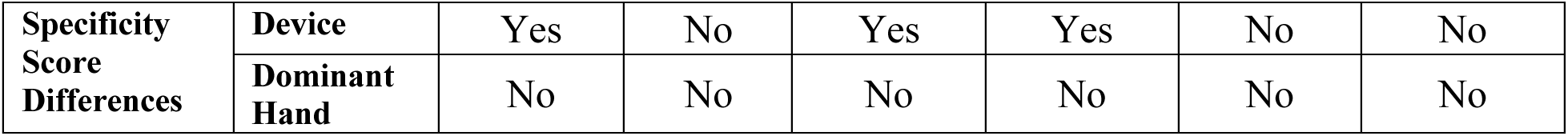
Binary indicator of whether a statistically significance difference in specificity is observed using a t-test between groups for each sensitive attribute (*before* race upsampling).

**Figure 4:**
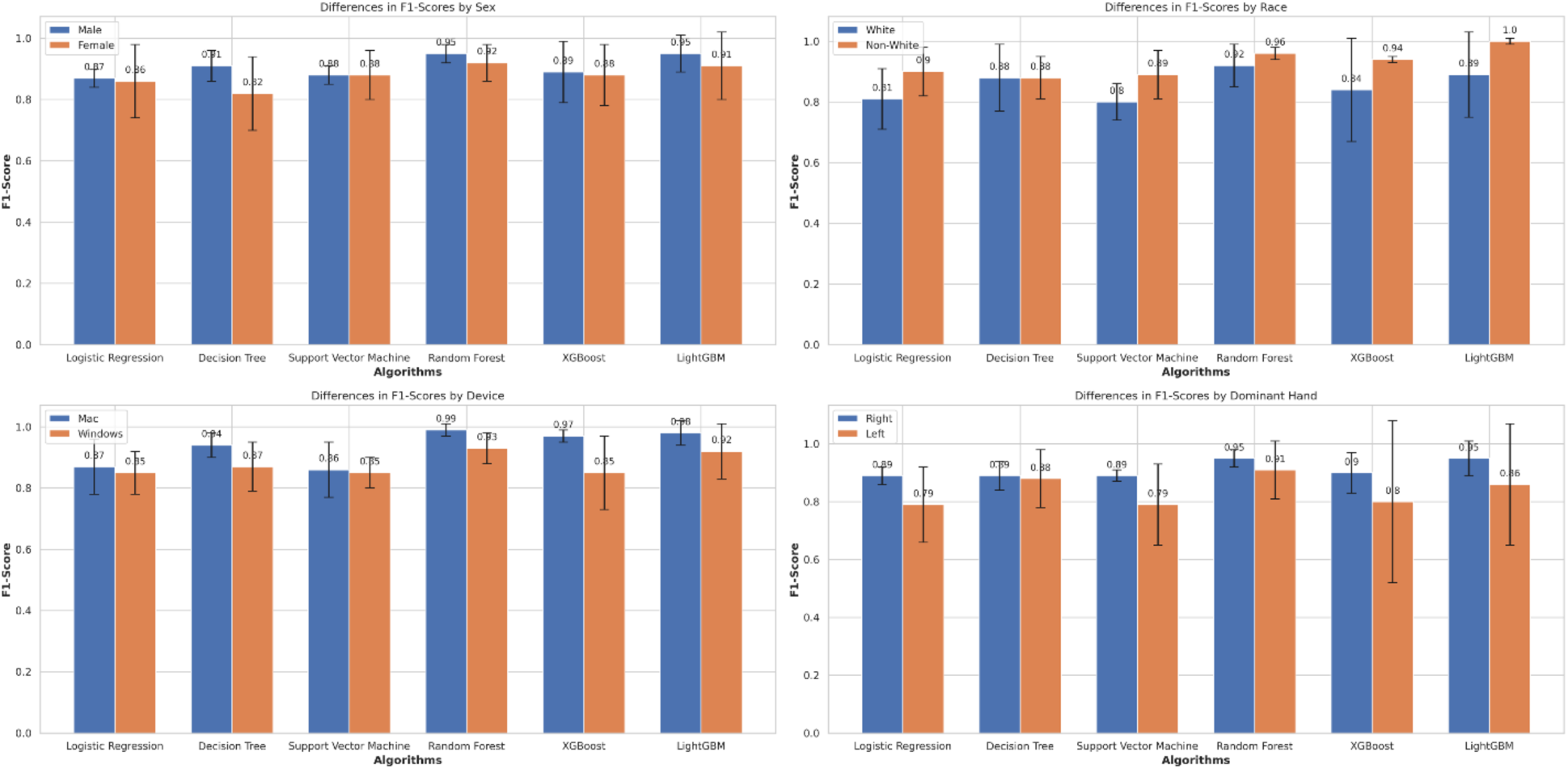
F1 score comparisons between the privileged and unprivileged groups for all protected attributes *after* race upsampling. No significant differences exist between the two groups in any category after upsampling by race.

Most algorithms achieved disparate impact scores somewhat close to 1 for sex and race after race resampling (Table 4.1). However, due to consistently high standard deviations, we cannot make any conclusive statements about whether sex or race significantly influences the models’ distribution of predictions towards any particular group.

Even after race upsampling, the models exhibit significant biases for two interesting attributes: device and dominant hand, with disparate impact values ranging from 0.38 to 0.70 and tighter standard deviations than for race or sex. This indicates that the unprivileged groups (Windows, left hand) are 38% to 70% less likely to receive a PD diagnosis from the model than the privileged groups (Mac, right hand).

## Discussion

Our PD assessment encompasses a novel approach to PD assessment. While prior PD studies have focused on speech data^4,18,19^, gait^8^, and handwriting^20^, our approach uniquely leverages data collected from a web application, enabling participants to contribute data remotely. This method aligns with the growing need for self-administered and non-invasive telemonitoring applications, which offer an accessible and convenient alternative to clinical examinations that are often challenging for elderly patients^21,22^. Moreover, prior studies have explored keyboard typing and touchscreen interactions for PD detection^23,24^, but few have addressed the issue of algorithmic biases^25^. These biases, often stemming from tiny sample sizes for underrepresented groups^26^, can compromise the fairness of machine learning models.

Although our models were generally performant, our analyses uncovered critical performance disparities, particularly concerning device type and dominant hand. Despite the goal of using our PD classification website to promote affordable and equitable screening, these biases highlight the need for quantifying biases – including non-obvious biases – in consumer digital health informatics.

Our results highlight the need for careful selection of AI bias metrics. Despite showing biases toward privileged groups in terms of receiving a PD diagnosis for device type and dominant hand, the models exhibited strong performance in terms of Equal Opportunity across all demographic groups, with values ranging between 0.87 and 1.22. However, this metric only captures the true positive rate (sensitivity) across groups and does not account for false positive rates or overall fairness. The Equalized Odds metric highlighted substantial imbalances.

In some cases, F1 scores were higher for one group than another, but AUROC showed the opposite trend. This discrepancy often occurs when a group’s specificity decreases significantly, affecting AUROC more than F1 scores. Conversely, there were instances where AUROC scores were higher despite lower F1 scores. In these cases, variations in precision mirrored those in F1 scores, while specificity and sensitivity remained strong, contributing to the higher AUROC values.

We note a few prominent limitations of our study. One significant limitation is that our model relied on data collected at a single point in time, thus failing to capture the progression of PD over time or symptomatic fluctuations. As PD is a degenerative condition, symptoms can vary due to factors such as medication, stress, and daily activities. Additionally, our study does not include detailed symptom quantification or information about participants’ medication usage or timing, critical factors that can significantly influence symptom severity and, thus, model predictions. We did not control for these factors. In future work, we aim to address these limitations by collecting longitudinal data and controlling medication timing and dose. In these future studies, it will be necessary to consider differences in symptom presentation variability across demographic groups, with the temporal aspect potentially introducing additional biases that were not explored in this work.

## Methods

### Study Design and Participant Recruitment

We recruited individuals aged 50 and above. The recruitment pool comprised both adults diagnosed with PD and healthy controls. To facilitate recruitment, we partnered with local organizations dedicated to PD support and advocacy, including through recruitment efforts at the 2023 and 2024 Hawaii Parkinson’s Association (HPA) Symposiums.

We invited participants to join the study using a web application accessible through a public link. The application provided detailed instructions about the research and its activities. Before commencing any activities, all participants provided informed consent electronically.

Figure 5 shows the overall procedure for collecting participant data, preprocessing the dataset to train the models, and evaluating the models for real-world application. The participants used the website to play different games, which required them to use the mouse to click and trace different objects and press keyboard buttons at various times during the game. These mouse trace data and keyboard press data were stored in a cloud database along with the participant’s demographic data (Figure 5a). The stored data were then analyzed and engineered to extract features from the mouse trace and keyboard press data. Finally, the dataset was split into 70% training and 30% testing data (Figure 5b). The dataset was trained using six common machine learning classification algorithms using 5-fold cross-validation, and the results were compared for further testing and evaluation on held-out data (Figure 5c). Afterward, the trained models were tested for bias in different demographic groups (Figure 6).

**Figure 5:**
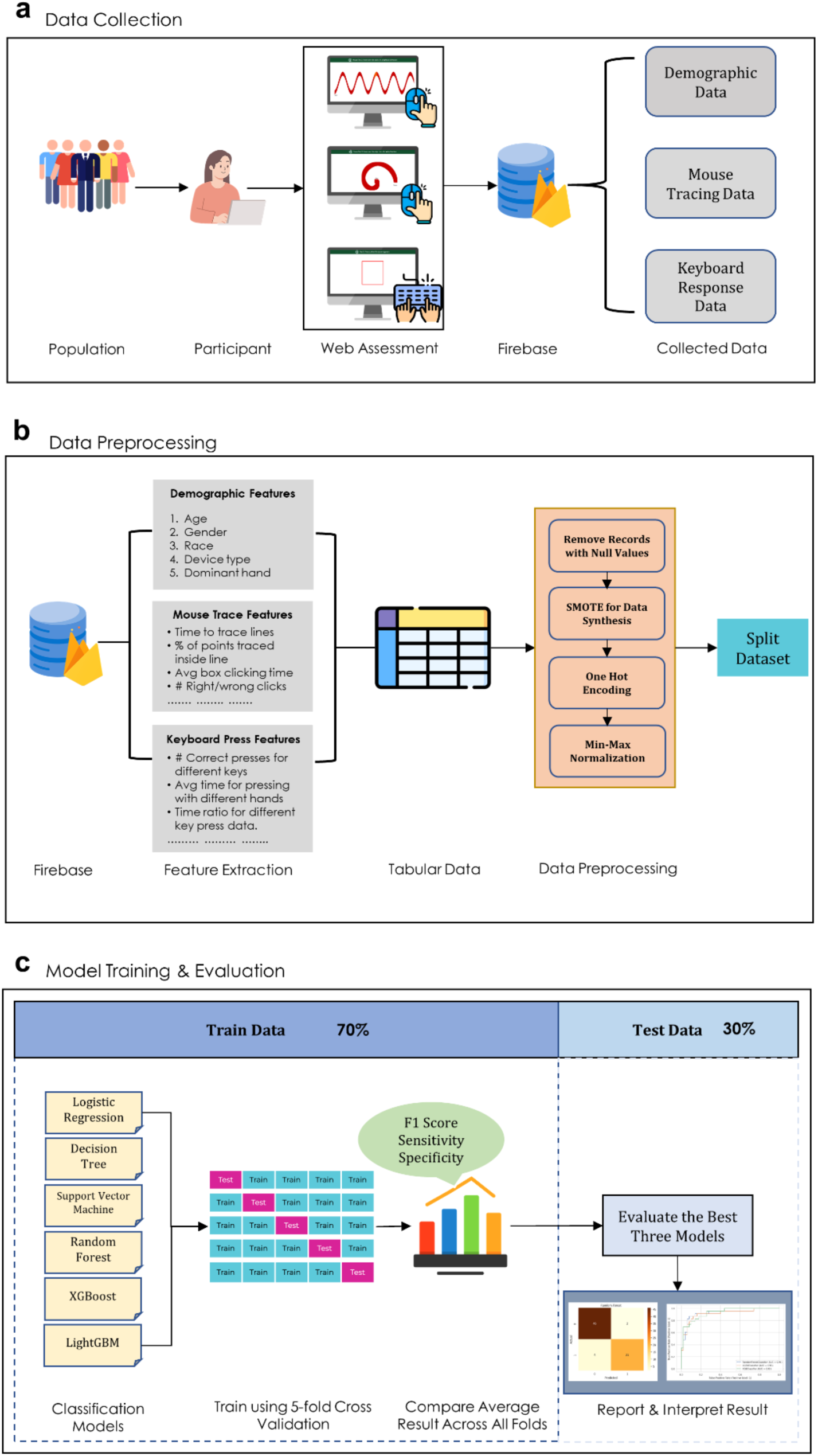
Study workflow. (a) We recorded hand movement data through a structured assessment on a publicly available website. The data are stored in a cloud database for further analysis. (b) Features are extracted from the hand movement data and organized into a tabular format. The features are preprocessed to remove any empty records. The categorical features were transformed using one-hot encoding, and numerical features were normalized using min-max scaling. (c) We trained the model with six machine learning algorithms using 5-fold cross-validation.

**Figure 6:**
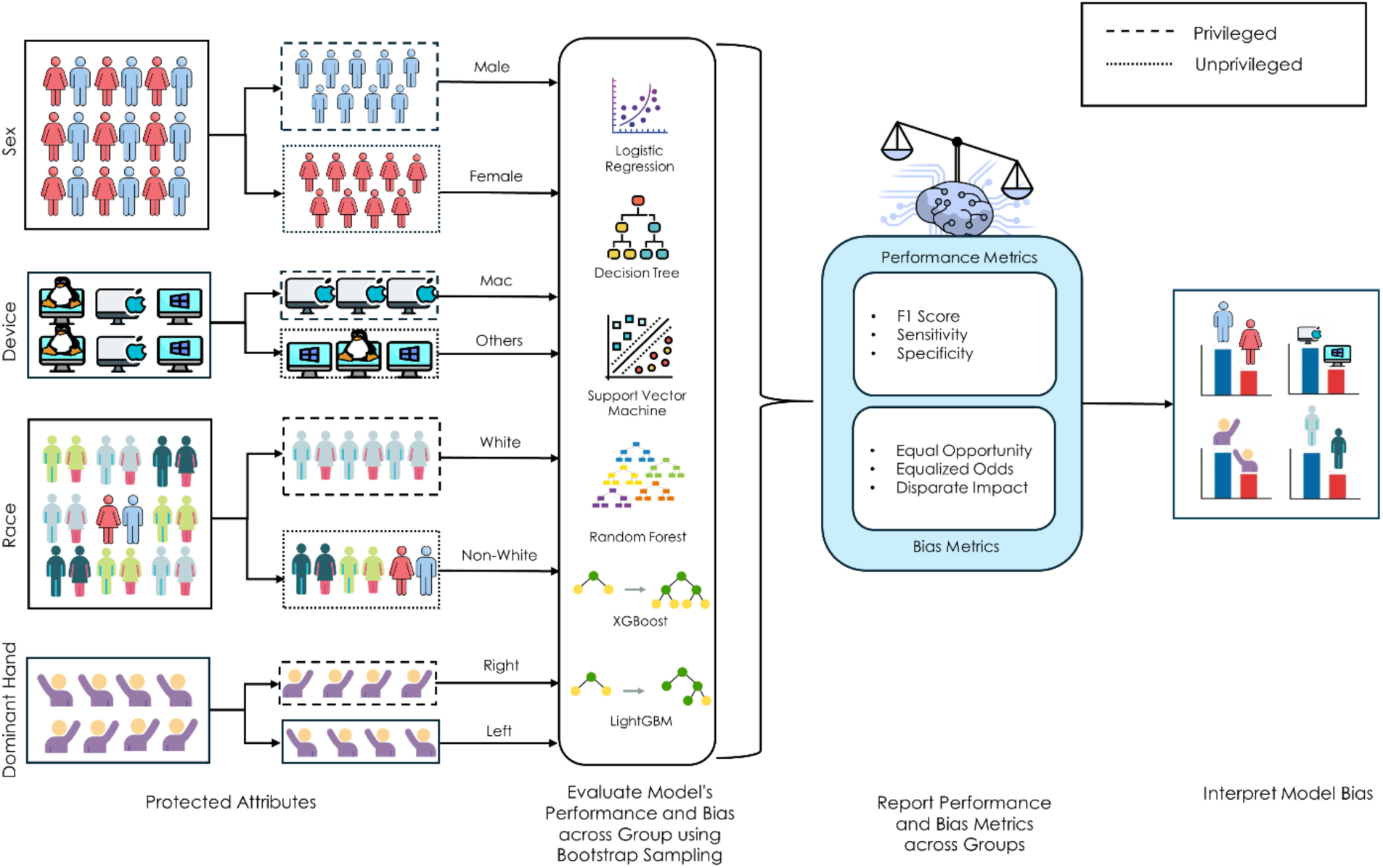
Bias evaluations. We quantified biases concerning sex, race, device type, and handedness.

### Data Collection Procedure

We recorded the participant data using a web-based assessment by evaluating motor functions (Figure 7). This evaluation was conducted through short and structured tasks designed to assess various aspects of user interactions.

**Figure 7:**
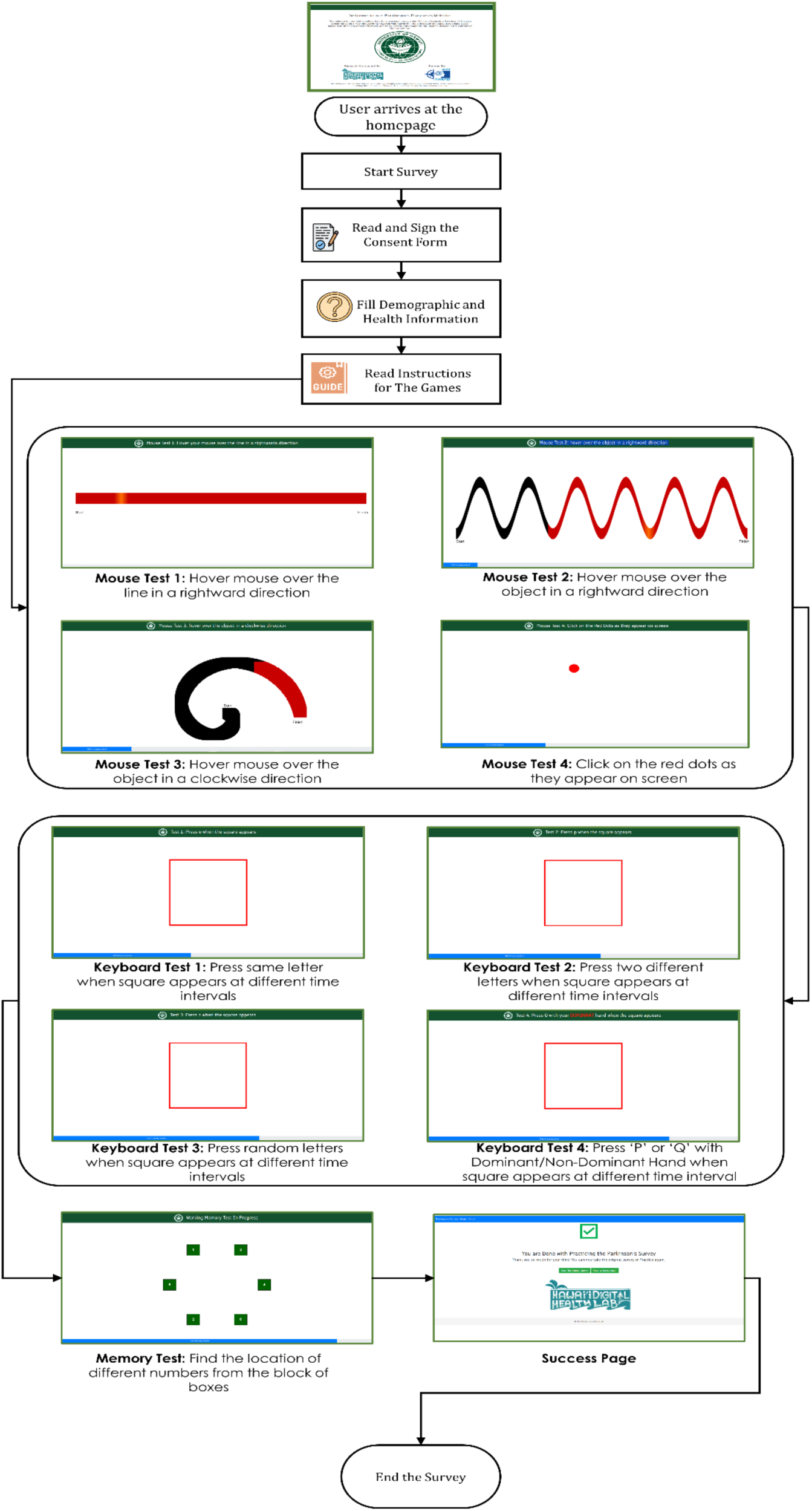
User interface of the web assessment. The user starts by filling out a consent form and records their demographic, health, and medication information. The users are then presented with shapes and required to trace the shape with their mouse/trackpad. This is followed by a series of games where the user is asked to press a key on the keyboard immediately when prompted. The game ends with a memory test we did not incorporate into our analysis.

The mouse-based tasks included:

- **Straight-Line Tracing**: Participants were tasked with tracing straight lines on the screen to evaluate hand stability and precision. This task required participants to maintain consistent control and accuracy, reflecting fine motor skills.
- **Curved Line Tracing**: Participants traced spirals and other curved shapes during this activity, offering a more complex evaluation of hand stability and motor coordination.
- **Randomized Clicking Tasks**: Participants clicked on targets appearing at random locations on the screen in order to measure reaction time, hand-eye coordination, and accuracy.

The keyboard-based tasks consisted of:

- **Single-Letter Presses**: Participants were instructed to press designated keys immediately upon receiving a visual prompt, assessing their motor response speed and accuracy.
- **Multiple-Letter Presses**: This task involved replicating key sequences, evaluating participants’ ability to execute complex motor tasks and process sequential information. In this phase, the task was confined to only two letters, and the user only needed to press one of the two letters that appear on the screen to pass.
- **Randomized Sequences**: Participants completed tasks involving sequences of randomized key presses, testing their adaptability, coordination, and cognitive flexibility.

We also added a Memory Game to assess cognitive function through progressively challenging levels. However, we did not use the cognitive assessment data in this study, and thus we do not include details about the assessment.

### Data Up-sampling

The initial dataset used in our study comprised 152 healthy individuals (60.6%) and 99 participants diagnosed with PD (39.4%). While the model initially achieved a strong F1 score, its sensitivity remained low, around 50%, which is particularly concerning in healthcare applications where detecting true positive cases is crucial. To address this issue and to improve the sensitivity of the models, we applied the Synthetic Minority Over-sampling Technique (SMOTE) to balance the dataset by up-sampling the PD patient data (updated data distribution shown in Figure 8).

**Figure 8:**
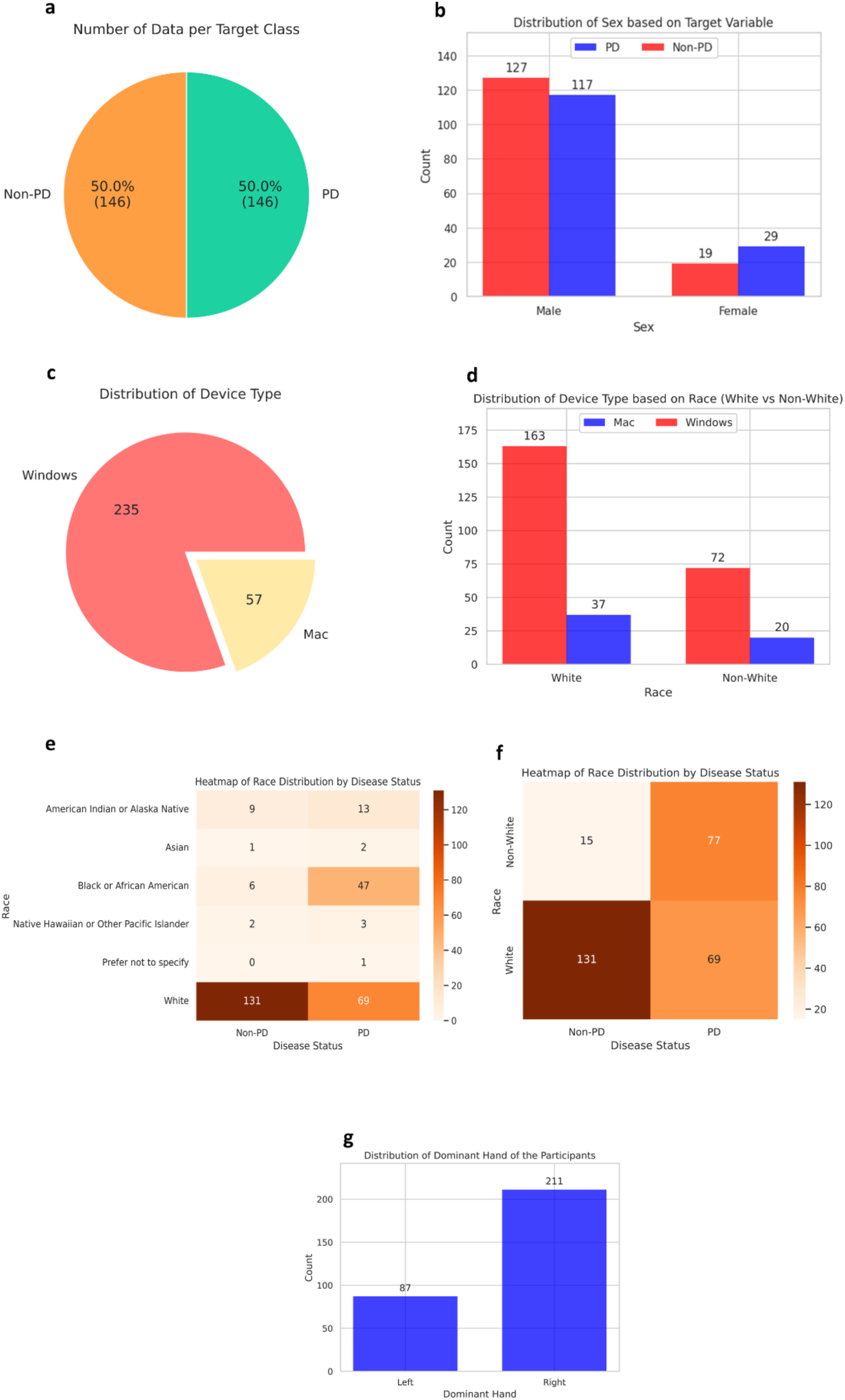
Data distribution after applying SMOTE.

Moreover, there was a significant imbalance in the racial distribution within the PD label, with 131 White participants compared to only 15 Non-White participants. To mitigate this imbalance, we applied a random resampling technique to increase the number of Non-White participants in the PD category (updated data distribution without SMOTE shown in Figure 9 and with SMOTE shown in Figure 10).

**Figure 9:**
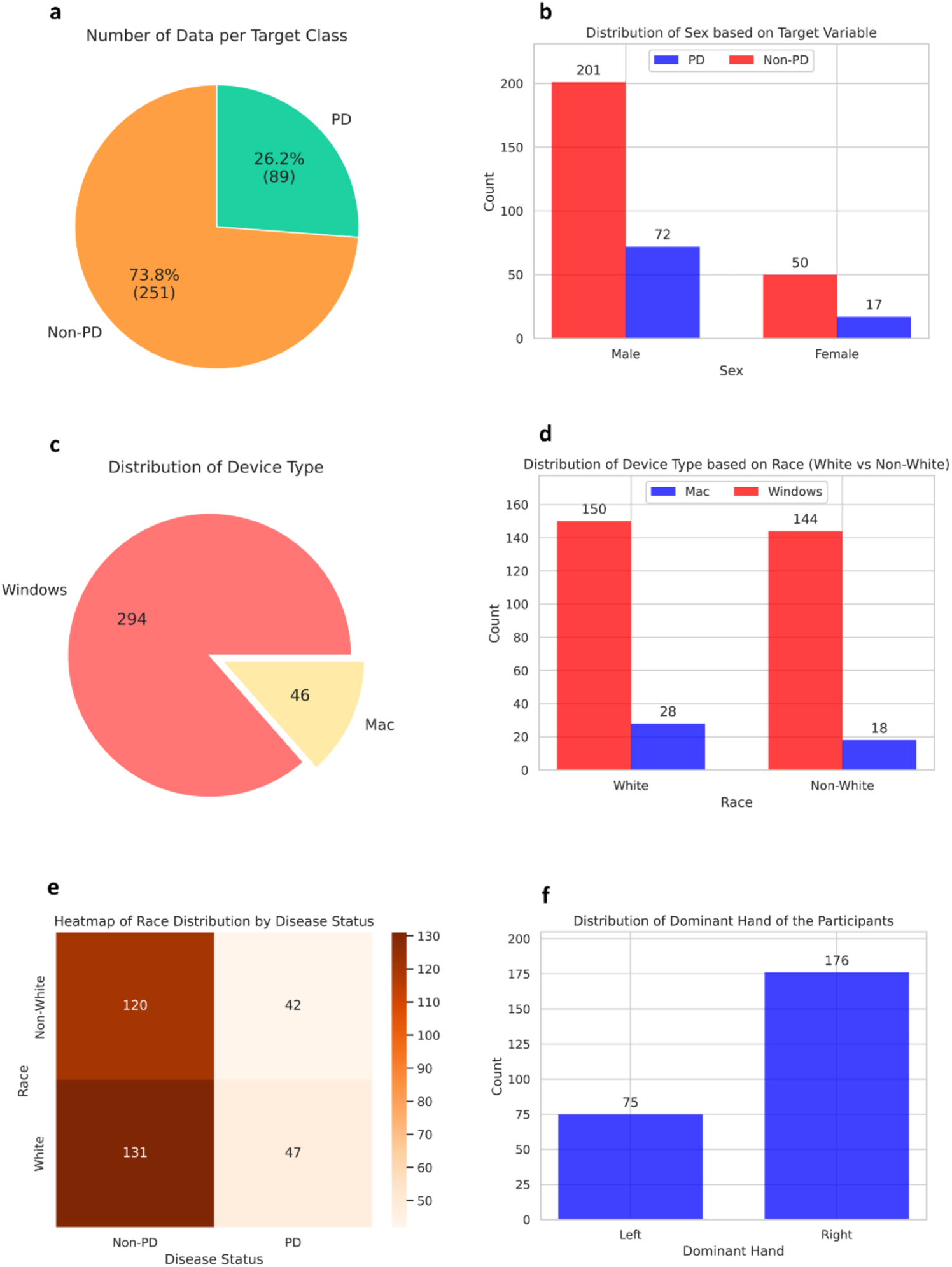
Data distribution after randomly resampling Non-White data points with no PD (before SMOTE).

**Figure 10:**
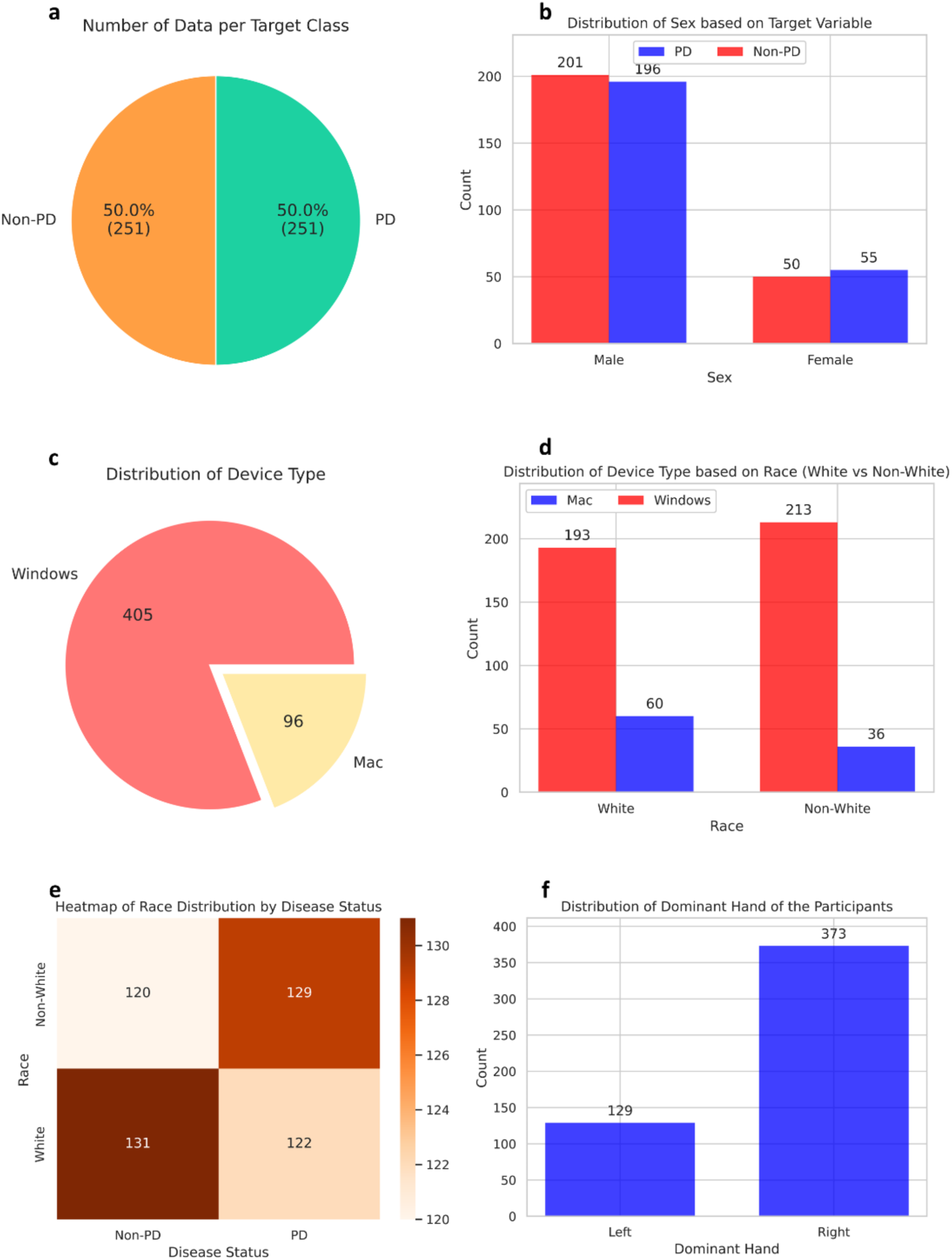
Data distribution after randomly resampling Non-White data points with no PD (after SMOTE).

### Bootstrapped Sampling

We apply bootstrapped sampling with 100 samples in each iteration to generate error bars. In each bootstrapped sampling step, the sample set consists of an equal number of data points from both groups in each category while calculating the group-wise metrics for that category, e.g., sex, race, device, and dominant hand. For example, when calculating the F1 scores, sensitivity, specificity, precision, and AUROC for sex, the sample set was constructed by taking 50% of the sample from the Male subset and 50% from the Female subset using resampling with replacement. Finally, we report the mean of the metrics.

### Feature Engineering

We engineered 80 features related to Hand Stability - Straight Line Tracing, Hand Stability - Curved Line Tracing, Response Times and Accuracy, Dominant Hand Movement, and Wrong Clicks, all engineered from the raw data. Table 6 summarizes some of the key engineered features we used in our study to detect PD.

**Table 6:**
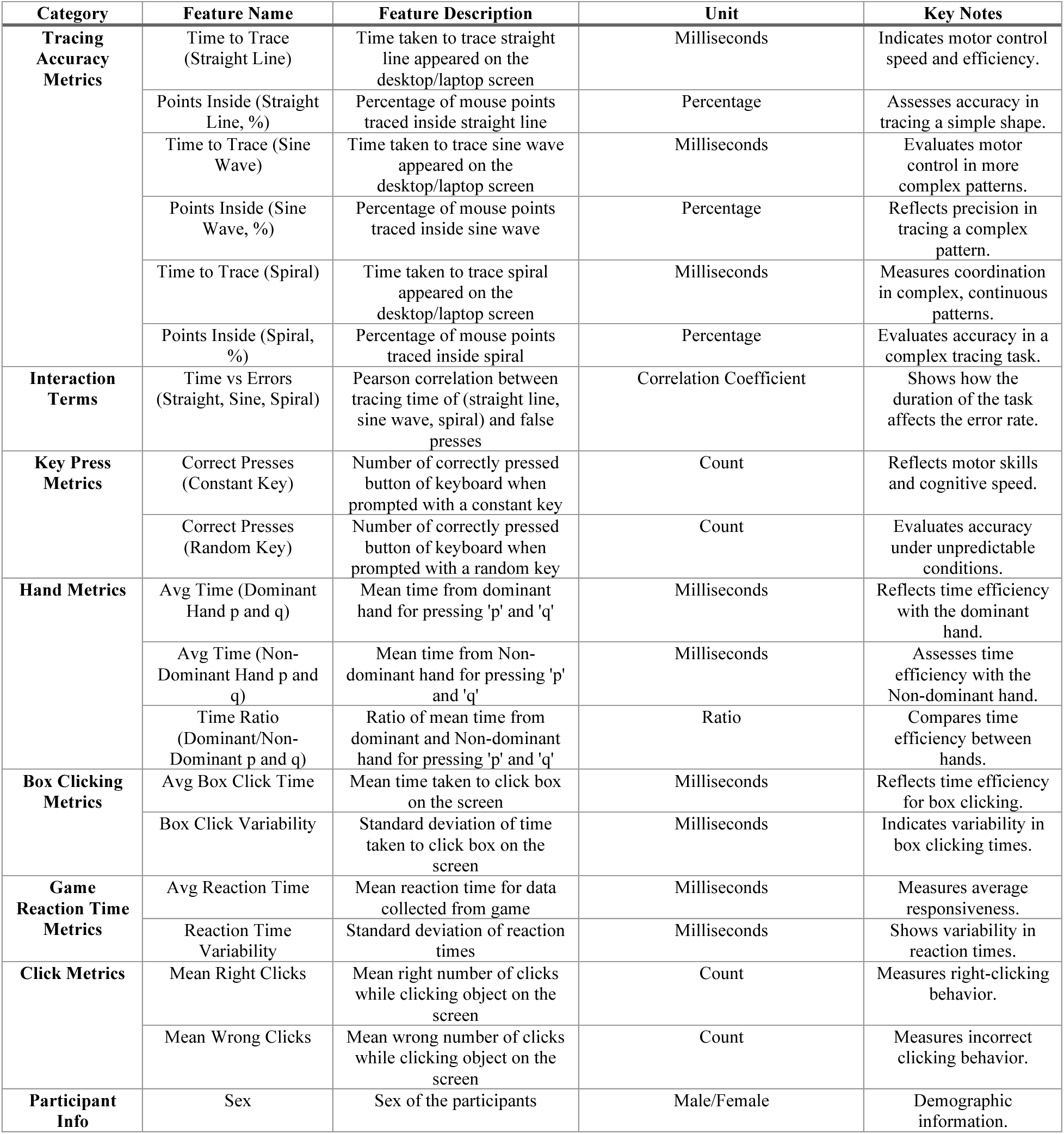
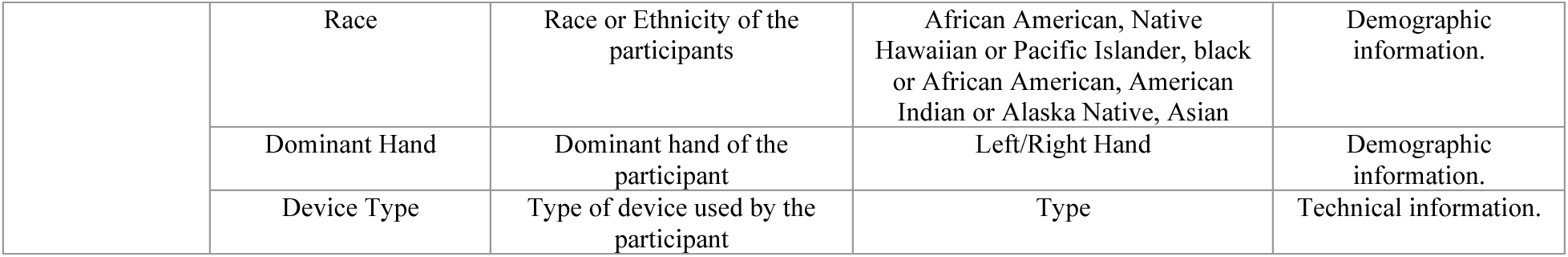
List of engineered features.

| Category | Feature Name | Feature Description | Unit | Key Notes |
| --- | --- | --- | --- | --- |
| **Tracing Accuracy Metrics** | Time to Trace (Straight Line) | Time taken to trace straight line appeared on the desktop/laptop screen | Milliseconds | Indicates motor control speed and efficiency. |
|  | Points Inside (Straight Line, %) | Percentage of mouse points traced inside straight line | Percentage | Assesses accuracy in tracing a simple shape. |
|  | Time to Trace (Sine Wave) | Time taken to trace sine wave appeared on the desktop/laptop screen | Milliseconds | Evaluates motor control in more complex patterns. |
|  | Points Inside (Sine Wave, %) | Percentage of mouse points traced inside sine wave | Percentage | Reflects precision in tracing a complex pattern. |
|  | Time to Trace (Spiral) | Time taken to trace spiral appeared on the desktop/laptop screen | Milliseconds | Measures coordination in complex, continuous patterns. |
|  | Points Inside (Spiral, %) | Percentage of mouse points traced inside spiral | Percentage | Evaluates accuracy in a complex tracing task. |
| **Interaction Terms** | Time vs Errors (Straight, Sine, Spiral) | Pearson correlation between tracing time of (straight line, sine wave, spiral) and false presses | Correlation Coefficient | Shows how the duration of the task affects the error rate. |
| **Key Press Metrics** | Correct Presses (Constant Key) | Number of correctly pressed button of keyboard when prompted with a constant key | Count | Reflects motor skills and cognitive speed. |
|  | Correct Presses (Random Key) | Number of correctly pressed button of keyboard when prompted with a random key | Count | Evaluates accuracy under unpredictable conditions. |
| **Hand Metrics** | Avg Time (Dominant Hand p and q) | Mean time from dominant hand for pressing 'p' and 'q' | Milliseconds | Reflects time efficiency with the dominant hand. |
|  | Avg Time (Non-Dominant Hand p and q) | Mean time from Non-dominant hand for pressing 'p' and 'q' | Milliseconds | Assesses time efficiency with the Non-dominant hand. |
|  | Time Ratio (Dominant/Non-Dominant p and q) | Ratio of mean time from dominant and Non-dominant hand for pressing 'p' and 'q' | Ratio | Compares time efficiency between hands. |
| **Box Clicking Metrics** | Avg Box Click Time | Mean time taken to click box on the screen | Milliseconds | Reflects time efficiency for box clicking. |
|  | Box Click Variability | Standard deviation of time taken to click box on the screen | Milliseconds | Indicates variability in box clicking times. |
| **Game Reaction Time Metrics** | Avg Reaction Time | Mean reaction time for data collected from game | Milliseconds | Measures average responsiveness. |
|  | Reaction Time Variability | Standard deviation of reaction times | Milliseconds | Shows variability in reaction times. |
| **Click Metrics** | Mean Right Clicks | Mean right number of clicks while clicking object on the screen | Count | Measures right-clicking behavior. |
|  | Mean Wrong Clicks | Mean wrong number of clicks while clicking object on the screen | Count | Measures incorrect clicking behavior. |
| **Participant Info** | Sex | Sex of the participants | Male/Female | Demographic information. |
|  | Race | Race or Ethnicity of the participants | African American, Native Hawaiian or Pacific Islander, black or African American, American Indian or Alaska Native, Asian | Demographic information. |
|  | Dominant Hand | Dominant hand of the participant | Left/Right Hand | Demographic information. |
|  | Device Type | Type of device used by the participant | Type | Technical information. |

### Evaluation Metrics

To evaluate the performance of our PD prediction model, we used a combination of standard performance metrics and fairness metrics to assess both accuracy and equity across demographic groups. This section provides a detailed description of these metrics.

### Standard Performance Metrics

We compute the F1-score, sensitivity (recall), precision, and specificity for each group across all demographic categories to evaluate model performance. Precision represents the proportion of correctly identified positive cases out of all predicted positive cases. Sensitivity (recall) measures the proportion of true positive (TP) cases accurately determined by the model. At the same time, specificity quantifies the proportion of true negative (TN) cases correctly classified, where true negatives refer to actual negatives correctly identified. In these calculations, false positives (FP) represent cases incorrectly classified as positive, and false negatives (FN) are actual positives misclassified as unfavorable. The F1-score combines precision and recall into a single metric, calculated as their weighted harmonic mean. The formulas for these metrics are:

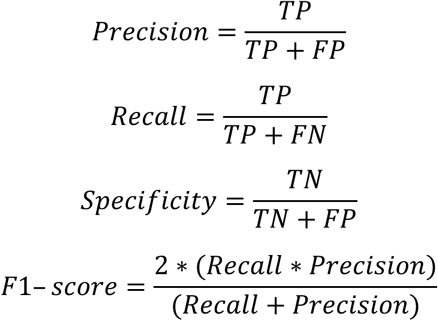

### Fairness Metrics

To ensure equitable outcomes across protected groups, we employed the following fairness metrics:

#### Disparate impact

Disparate Impact (DI) is a fairness metric used to evaluate equity in model outcomes by comparing the rates of positive outcomes between a privileged and an unprivileged group ^29,30^. It assesses whether the unprivileged group is disproportionately less likely to receive favorable outcomes than the privileged group. DI is calculated as the ratio of the proportion of positive outcomes for the unprivileged group to that of the privileged group, defined as:

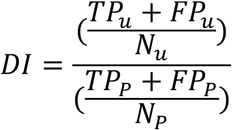

*TP_u_* and *TP_p_* represent true positives while *FP_u_* and *FP_p_* denote false positives for the unprivileged (u) and privileged (p) groups. *N_u_* and *N_p_* denote the total number of individuals in the unprivileged and privileged groups, respectively.

To achieve fairness, the Disparate Impact (DI) value must equal 1, indicating equal rates of positive predictions for both groups^29^. A value below 1 signals potential bias against the unprivileged group, while a value above 1 suggests a potential bias favoring the unprivileged group.

#### Equal Opportunity

Equal Opportunity (EO) is a fairness concept requiring that privileged and unprivileged groups achieve equal true positive rates (TPR)^31^. This ensures that individuals from all demographic groups who qualify for a positive outcome are equally likely to receive it. EO is mathematically defined as the ratio of the TPRs between unprivileged and privileged groups and is expressed as:

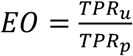

An Equal Opportunity (EO) value of 1 represents perfect fairness, signifying that true positive rate (TPR) are identical across both privileged and unprivileged groups. A value less than 1 indicates that the unprivileged group has a lower TPR than the privileged group, suggesting potential bias against the unprivileged group in receiving favorable outcomes. Conversely, a value greater than 1 indicates that the unprivileged group has a higher TPR than the privileged group, which may reflect a bias favoring the unprivileged group.

#### Equalized Odds

Equalized Odds require that both true positive rates (TPRs) and false positive rates (FPRs) are equal across privileged and unprivileged groups^31^, ensuring that the model’s predictions regarding successes and errors are fair. By extending the concept of Equal Opportunity, Equalized Odds ensures that predictions do not disproportionately favor or disadvantage any group. It can be mathematically represented as the ratio of TPRs and FPRs between the unprivileged and privileged groups:

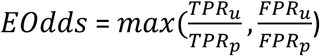

An Equalized Odds value of 1 represents ideal fairness, signifying uniform error distribution and success rates across all demographic groups. Deviations from this standard, whether below or above 1, indicate disparities in the model’s predictions, pointing to potential biases in true positive rates (TPRs), false positive rates (FPRs), or both.

## Data Availability

All data produced in the present study are available upon reasonable request to the authors.

## Ethical Considerations

This project received approval from the University of Hawaii Institutional Review Board (IRB), protocol #2022-00857.

## Acknowledgments

This research was partly funded by the National Institutes of Health (NIH) Agreement No. 1OT2OD032581-01. The views and conclusions in this document are those of the authors and should not be interpreted as representing the NIH’s official policies, either expressed or implied. The language and grammar of this manuscript were revised using AI tools such as ChatGPT and Grammarly. However, the authors thoroughly refined the language after the stylistic revisions provided by these tools.

